# Monitoring and predicting viral dynamics in SARS-CoV-2-infected Patients

**DOI:** 10.1101/2020.04.14.20060491

**Authors:** Shaoqing Wen, Yi Wang, Jianxue Xiong, Chang Sun, Barnaby Edward Young, David Chien Lye, Yee Sin Leo, Li Jin, Guochang Wang

**Author notes:** These authors contributed equally to this work (Shaoqing Wen, Yi Wang).

## Abstract

This study is based on the a simple but robust model we developed urgently to accurately monitor and predict viral dynamics for each SARS-CoV-2-infected patient, given the limited number of RT-PCR tests and the complexity of each individual’s physical health situation. We used the estimated regression model to monitor and predict the changes of viral loads from different nasal and throat swab of clinical specimens collected from diagnosed patients. We also tested this real-time model by using the data from the SARS-CoV-2-infected patients with different severity. By using this model, we can predict the viral dynamics of patients, minimize false-negative test results, and screen the patients who are at risk of testing positive again after recovery. We sincerely thank those who are on the front lines battling SARS-CoV-2 virus. We hope this model will be useful for SARS-CoV-2-infected patients.

---

Due to the principle of early identification, early isolation, early diagnosis and early treatment, the 2019 novel coronavirus (SARS-CoV-2) epidemic in China is under control. Viral dynamics studies are an important source of information on SARS-CoV-2 pathogenesis. The latest studies about SARS-CoV-2 viral dynamics revealed that: 1) the viral load in lower respiratory tract (LRT; such as sputum) is generally higher than upper respiratory tract (URT; such as nasopharyngeal and oropharyngeal) [1] and, in the upper respiratory tract, higher viral loads are detected in the nose rather than in the throat [2]; 2) the viral loads in SARS-CoV-2-infected patients peaked soon after the onset of symptoms, resembling influenza-infected patients and distinct from SARS-CoV-infected patients [1-4]; 3) viral loads detected in asymptomatic or minimally symptomatic patients indicate the transmission potential in the early phase of infection [2].

However, there are many details that remain puzzled. For example, 1) some patients have presented positive chest CT findings showing multifocal ground-glass changes, but they had negative RT-PCR results at that time [5]; 2) some patients have rapid changes in viral loads during the window of infection, with the RT-PCR results sometimes positive and sometimes negative [2-4]; 3) few recovered patients (absence of clinical symptoms and radiological abnormalities and 2 consecutively negative RT-PCR test results separated by at least 1 day) had positive RT-PCR test results again 1 or 2 weeks after discharge from hospital [6]. Importantly, it is impossible to continuously collect samples from each SARS-CoV-2-infected patient over the course of the treatment and rehabilitation.

In this context, a simple but robust model is expected to accurately monitor and predict viral dynamics for each SARS-CoV-2-infected patient, given the limited number of RT-PCR tests and the complexity of each individual’s physical health situation.

First of all, we reformat the original sparse matrix drawn from ref [2] (viral loads of 18 Chinese infected patients with many missing data points) into a dense matrix (training set) as follow:

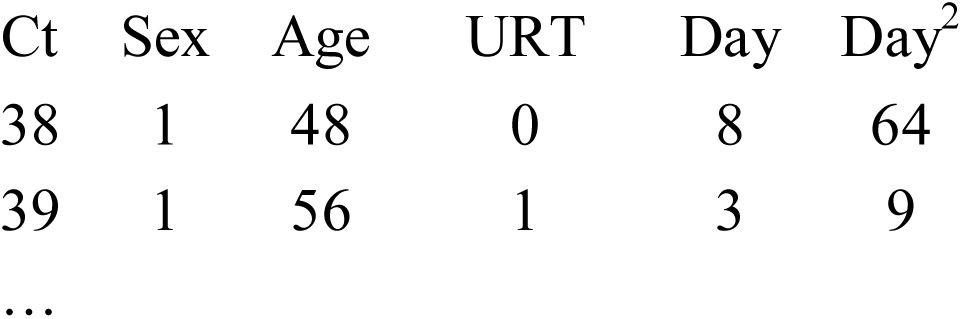

We use a linear mixture model to predict the Cycle threshold (Ct). Ct values of SARS-CoV-2-specific gene on RT-PCR assay are inversely related to viral RNA copy number. Note that nasal data and throat data are also combined with an indicator URT in order to maximize the number of data points. The specific linear mixture model is defined as

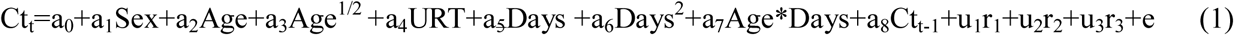

where Age*Days is the interaction term of Age and Days; Ct_t_ and Ct_t-1_ are the Ct values of the t-th and (t-1)-th day; u_1_=1 if the symptom of the patient is mild, otherwise u_1_=0, and u_2_, u_3_ are defined similarly, u_2_=1 if the symptom of the patient is moderate, and u_3_= 1 if the symptom of the patient is severe; Sex, Age, …, Days^2^ and Age *Days are the fixed term; a_0_, …, a_8_ are the corresponding coefficient; u_1_, u_2_, u_3_ are also observed, but r_1_, r_2_, r_3_ are random variable and called it random effect. To simple the compute, we usually assume that r_i_ is normal distribution with mean zero and variance is 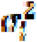. We refer the readers to Wang et al. [7] for the specific estimate method. We can not build too complex statistical model since we only have 18 samples. To solve this problem, we do not build a global model for each patient, but build a model for each day after onset. And this leads to one patient has more than one observation. For an example, if one patient has five Ct values after onset, then, it is equivalently to have four observations or samples. And the total sample size for model (1) is 90, which is enough to estimate the model coefficient and the random effect term. We use all of 90 samples to estimate the random effect term, which can enhance the performance of prediction. The coefficient a_0_, …, a_8_, the testing for coefficient and some other model information including the R-square, the model F-statistics and P-values are all summarized in Supplementary Table 1.

Subsequently, we tested the personal model with another two sets of published data [4,5]. The test set included 13 mild patients, 5 moderate patients (requiring supplemental oxygen), and 2 severe patients (admitted to ICU), of which 7 patients were treated with lopinavir-ritonavir. The detailed epidemiologic features and clinical course of these patients are described in Supplementary Table 2. The Ct values of 20 confirmed patients can be also found in Supplementary Table 2, according to the timeline from symptom onset. For each SARS-CoV-2-infected patient, we predicted the Ct values in the near future based on the tested Ct values (Figure 1 and Supplementary Table 2). Figure 1 presented the Ct value curves of the 20 confirmed cases within 21 days since onset of symptoms, because of the time restriction of training set [2]. The absolute value of errors between predicted and tested Ct values is also shown in Supplementary Table 2. Over all, the mean and median of absolute value of errors is 2.99 and 2.82, respectively. The mild, moderate and severe patient group gave the average absolute value of errors for each patient, ranging from 1.33-7.00 (mean 3.08, median 2.96), 1.88-1.95 (mean 2.01, median 1.95), and 2.79-3.76 (mean 3.28, median 3.28), respectively. There was no significant difference between the drug group (mean 2.93, median 2.70) and non-drug group (mean 2.72, median 2.30). In figure 1, we observed that most samples’ predicted Ct value peak at around 5-6 days after symptom onset, which is consistent with previous studies [1]. However, for each SARS-CoV-2-infected patient, the case ending time is quiet different. Within 21 days after symptom onset, we predicted that 14 patients’ viral loads would become undetectable. Notably, we did observe the drastic fluctuations of predicted Ct value curves. Some patients (such as patient 7, 8, 10, 11, 14, 16, and 17) have the positive and negative RT-PCR results, alternately. Combined with our predicted Ct value curves, that could be because the treatment with lopinavir-ritonavir, or, more likely, because the occurrence of false-negative test results. Moreover, we should pay special attention to those who have negative RT-PCR results but lower predicted Ct values on day 21 after symptom onset, such as patient 7 and 11, which implies they may turn positive again.

**Figure 1.**
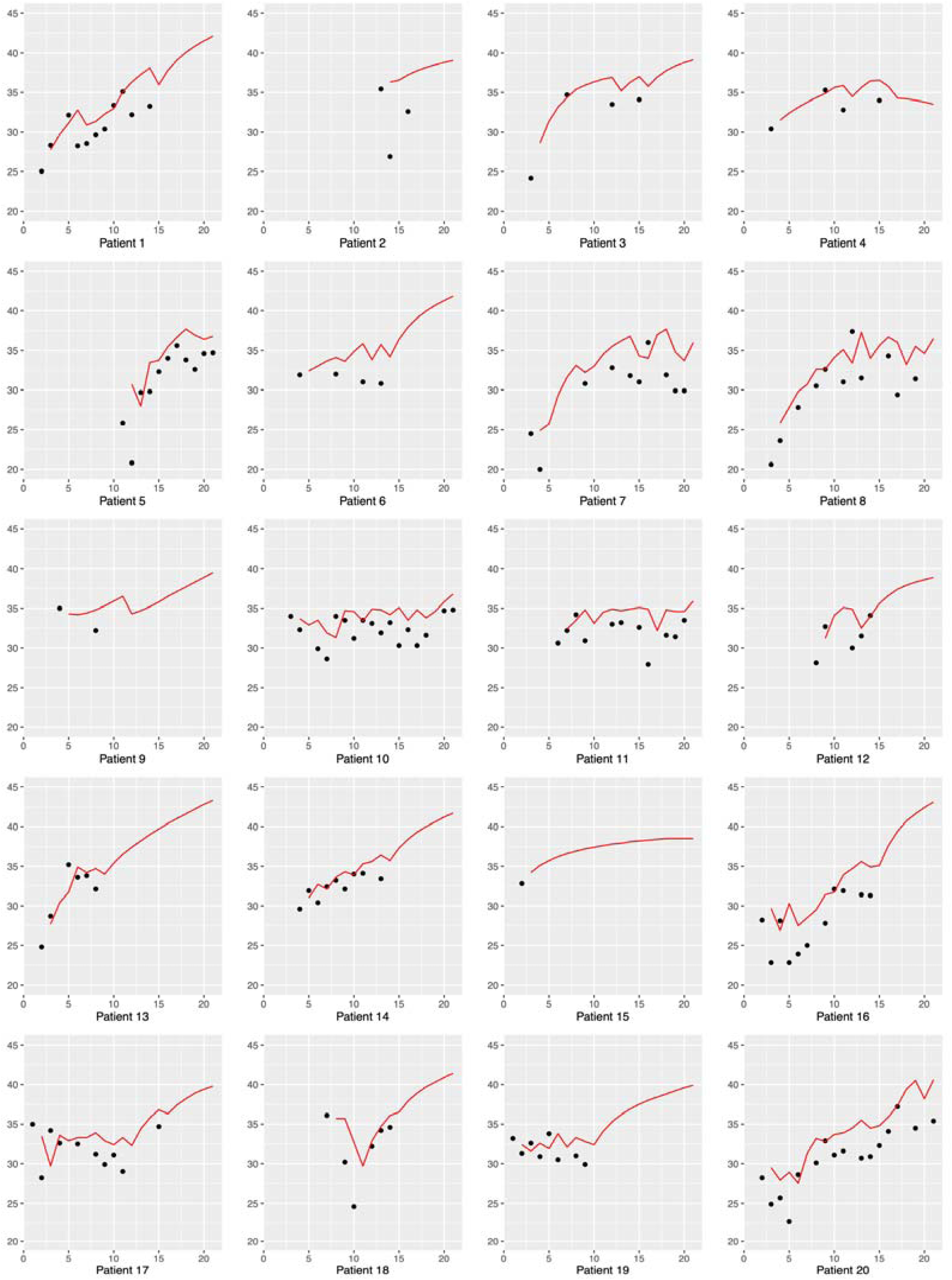
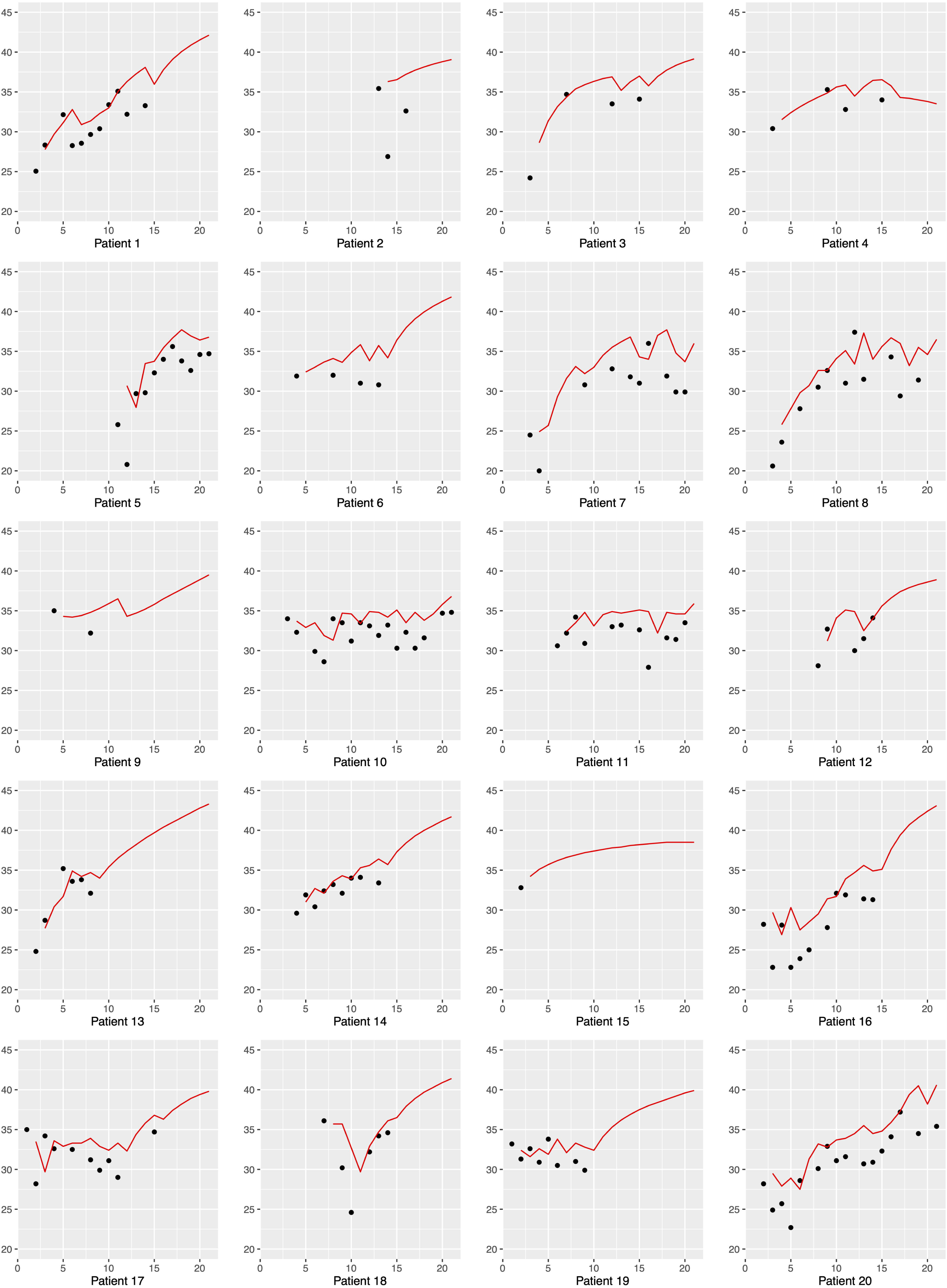
The tested Ct values of 20 SARS-CoV-2-infected patients and their predicted Ct value curves.

In this study, we used a mathematical model to monitor and predict the changes of viral loads from different nasal and throat swab of clinical specimens collected from diagnosed patients. We also tested our real time model by using the data from the SARS-CoV-2-infected patients with different severity. By using this personal model, we can predict the viral dynamics of patients, minimize false-negative test results, and screen the patients who are at risk of testing positive again after recovery. The following factors can optimize our model:

1. more data regarding viral loads of the infected patients;
2. detailed information about clinical features and laboratory results;
3. more samples covering the clinical course, especially key time points (exposure, symptom onset, 5-6 days after symptom onset, recovery, and two weeks after recovery);
4. more attention to the early phase between exposure and symptom onset, which is currently a knowledge gap for SARS-CoV-2 viral dynamics studies.

Finally, we hope this model will be useful for SARS-CoV-2-infected patients.

## Data Availability

All data can be obtained via the supplementary materials of the formally published paper.

http://58.87.113.187:8080/

## Supporting Information

Table S1. The basic information of the estimated regression model in this study.

Table S2. Clinical course and viral loads (tested Ct values and predicted Ct values) of 20 SARS-CoV-2-infected patients according to the timeline from symptom onset.

## Acknowledgments

We sincerely thank those who are on the front lines battling SARS-CoV-2 virus. The method can be realized by SPSS or R software. Groups interested in testing the model can request the code by emailing.

