## Supplemental Table 1 for "Monitoring and predicting viral dynamics in SARS-CoV-2-infected Patients"

Table 1£º the estimated regression model
Variable         coefficient           t-statistic            p-value                

Intercept         23.95879              0.03160          0.48743
Sex              0.01422               0.01213          0.49518
Age             0.34552               0.62615          0.26649
Age^1/2^           -4.07916              -0.04928          0.51959
URT             0.56162               0.56410          0.28712
Days             0.53289              0.89496          0.18673
Days^2^             -0.00391             -6.69141         1.00000
Age*Days         -0.00525             -20.98257        1.00000
Ct_t-1_                          0.59131               46.14499         0.00000^***^

RMSD for the training sample:  4.309979£¬   R-Squared : 0.384201
RMSD for the testing sample: 3.392093
F-statistic: 50.5155 on 1 and 81 degree Freedom, p-value: 4.180047e-10^***^
               
	
Signif. codes: 0.01 '***', 0.05 '**', 0.1 '*' ,   , where  is the estimated values and  is the true value. 
